# Cutaneous Leishmaniasis in Tigray, North Ethiopia: The Communities’ Awareness, Perceptions, Treatment-seeking and Prevention Practices in Disease Endemic Areas

**DOI:** 10.64898/2026.03.26.26349367

**Authors:** Shewaye Belay Tessema, Helen P Price, Afework Mulugeta Bezabih

## Abstract

**Background:** Cutaneous leishmaniasis (CL) is highly prevalent in Ethiopia, including the Tigray region. However, there is a dearth of information on the levels of knowledge, attitude, and health seeking behavior among the communities in CL-endemic areas of Tigray region, northern Ethiopia.

**Objective:** This study aimed to investigate CL-related knowledge, attitude, treatment-seeking and prevention practices in disease-endemic areas of Tigray.

**Methods:** Between November and December 2022, a cross-sectional survey was conducted among communities living in seven districts of Tigray. A mixed sampling method was implemented. Data were collected using a pre-tested structured questionnaire and analyzed using SPSS 25 (IBM, Chicago).

**Results:** A total of 512 participants were included. Overall, 43%, 36% and 34% of participants had a ‘good’ level of knowledge, a ‘favorable’ attitude and a good treatment-seeking and prevention practices towards CL, respectively. However, nearly all participants did not know about CL transmission, about 25% perceived CL to be genetically acquired and about 67% believed it to be stigmatizing. Traditional medication was the preferred option over modern treatment for 63.3%. Rural dwelling participants (AOR = 1.60; 95% CI: 1.00–2.57) and participants living in households with CL episode (AOR = 10.19; 95% CI: 6.36–16.30) had good knowledge towards the disease. However, urban/ semi-urban residents (AOR = 2.17; 95% CI: 1.42–3.31) had favorable attitude towards CL. Gender (AOR = 1.49; 95% CI: 1.01–2.22) and education level (AOR = 0.39; 95% CI: 0.24–0.62) were significantly associated with treatment-seeking and prevention practices. Participants living in households with CL episode (AOR = 2.99; 95% CI: 1.96–4.57) had good treatment-seeking and prevention practices.

**Conclusion:** In this study, over one half of participants had poor knowledge about CL, nearly two-third of them had unfavorable attitude towards the disease and two-third of them had poor treatment-seeking and prevention practices. Residence and previous CL episode in households were determinants of respondents’ knowledge about CL and their attitude towards the disease. Level of education and living in households with CL episode were determinants of participants’ treatment-seeking and prevention practices. These findings support for an integrated intervention through health education focusing on CL transmission and preventive measures.

**Authors’ summary:** Cutaneous leishmaniasis (CL) is one of the major health challenges in the highlands of Tigray, Northern Ethiopia. However, little is known about the communities’ awareness and prevention practices in the disease endemic areas of the region. In this study, we evaluated the knowledge, attitude, practice (KAP) and treatment seeking behaviors and prevention practices of communities located in seven CL-endemic districts of Tigray region. Our findings indicate that most of participants (96.8%) had seen CL cases within the study communities, either from infected household family or neighbors, and considered a skin lesion on the face to be a symptom of CL. However, only 43% of participants had an overall ‘good’ level of knowledge. Moreover, they didn’t know about the mechanisms of CL transmission, only six participants (1.2%) mentioned the sand fly vector by name and about 25% still perceived CL to be genetically acquired. Furthermore, more than two-third of participants had unfavorable attitude towards the disease and over 67% perceived CL to be a stigmatizing disease. Over two-third of the current participants had a poor level of treatment-seeking behaviour, over 63% preferred to seek remedies from traditional healers rather than formal CL healthcare facilities. Besides, over 71% of respondents didn’t apply any known preventive measures of CL and over 66% had a poor level of prevention practices. Occupation and rural household settings were determinants significantly associated with participants’ attitude towards CL. Gender and previous CL episode in households were also significant predictors associated with treatment seeking behavior and prevention practices. These findings highlighted for an integrated intervention through community mobilization and education campaigns focusing on CL causal agents, transmission mechanisms and its presentation measures.

## Introduction

Cutaneous leishmaniasis (CL), one of the vector-borne neglected tropical disease (NTDs), is caused by protozoan parasites of the genus *Leishmania* and transmitted via the bites of infected female sandflies of over 90 species [1]. The disease is endemic in more than 90 countries worldwide. There are approximately 0.7 to 1.2 million new cases annually across the globe and around 350-430 million people are estimated to be at risk of infection [2–5]. Furthermore, about 40 million people globally are living with CL scars from past infections [6], which is considered to be a leading cause of stigma, depression and anxiety among NTDs [7]. However, there is substantial under reporting of this disease, with only 200,000 cases per annum being reported to the World Health Organization (WHO) [3, 8, 9].

In Ethiopia, the report of CL dates back at least to the early 1900s, with the first case reported from the northern part of the country and described as “Oriental sore in Agamé (Abyssinia)” [10]. Since then, the disease has been reported from highlands of the country and over 170 districts are suspected to be endemic [11]. In Ethiopia, CL presents in three clinical forms: localized (LCL), diffused (DCL) and mucocutaneous (MCL) [12] of which MCL and DCL present significant therapeutic challenges [13]. Studies have estimated that 20,000 to 50,000 people per year are being infected in Ethiopia [2, 14]. However, in 2022, only 913 CL cases were reported to the WHO [8] suggesting a gross underreporting of the disease in Ethiopia.

An effective way to alleviate the burden of infectious diseases is to improve the communities’ awareness and prevention practices [9, 15]. Understanding the perceptions and behaviors of the affected community towards CL is valuable for designing control and prevention strategies for the disease [16]. The knowledge, attitude, practice (KAP) evaluation surveys are basic inputs for health promotion campaigns, as the surveys help programs adjust health education messages to build public knowledge and awareness [17]. Several studies indicate a direct relationship between community awareness on CL and effectiveness of control strategies [18, 19]. Earlier study reports implicate that the northern part of Ethiopia bears the highest burden of CL relative to the rest part of the country [14, 21]. The knowledge, attitude, practice (KAP) and treatment seeking behaviors and prevention practices towards CL in Ethiopia are heavily influenced by socio-cultural settings and widely vary among communities across different regions of the country, however, only a small number of KAP studies have been conducted in CL-endemic regions [20–22]. Moreover, community health education towards prevention and control of the disease in Ethiopia is lacking [12]. Furthermore, in the Tigray region, information related to CL endemic communities’ KAP and treatment-seeking behavior are very scarce. Therefore, in the present study, we evaluated the knowledge, attitude, practice (KAP) and treatment seeking behaviors and prevention practices of communities located in seven CL-endemic districts of Tigray region, northern Ethiopia.

## Methods

### Study setting

The Tigray region is located in the Northern part of Ethiopia between 12° 15‘N and 14° 57‘N latitude and 36° 27‘E and 39° 59’E longitude. The Tigray region has 7 administrative zones namely Central, Eastern, Mekelle, North Western, Southern, South Eastern, Western and Mekelle; 52 districts (locally known as *woredas*); and 799 sub-districts or *kebeles* (locally, *Tabias*). A *Tabia* is the smallest administrative unit in the region which consists of 8 to 10 small villages (called *kushets*) and a *kushet* comprises on average from 150 to 250 households. The topography of Tigray consists of high plateau and mountains with much of the land lying between 1,000 and 3,900 meters above sea level altitude. This study was conducted in seven CL endemic districts found in three zones of Tigray, namely: *Ganta Afeshum, Gulomekeda, Hawzen* and *Saesie Tsaeda-emba* located in Eastern zone, *Degua Temben* and *Enderta* in South Eastern, and *Emba-alaje* situated in Southern zone.

### Study design and study period

A cross-sectional survey was carried out between November and December 2022, in seven districts located in three zones of Tigray, Northern Ethiopia.

### Sample size

The minimum sample size required for this survey was calculated according to the WHO’s practical manual for sample size determination in health studies [23]. As previous data towards the level of community awareness and practices related to CL in the study areas were unavailable, we assumed 50% level of awareness and practices in the targeted area study communities. The sample size was calculated using a single population proportion formula, assuming 95% CI with 0.05 margin of error and a 1.3 design effect.

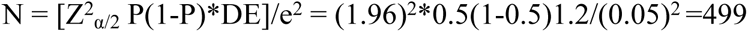

The minimum sample size required for this study was yielded 499 respondents using the following assumption: N = the number of study subjects (household heads), Z is a critical value (1.96) at 95% confidence level, P = anticipated population proportion (50%), DE = design effect and e= margin of error (5%).

To the calculated minimum sample size (n=499), a 5% of non-response rate was added and the total sample size was determined to be 524 individual participants.

### Sampling procedure

Between March and May 2022, we have conducted CL epidemiological baseline surveys (Tessema *et al*., 2026; https://doi.org/10.1371) in seven districts located in three zones of Tigray by hosting 3,817 participants resided in 927 households [24]. The districts included in the baseline CL survey were: *Ganta Afeshum, Gulomekeda, Hawzen* and *Saesie Tsaeda-emba* districts (from in Eastern zone), *Degua Temben* and *Enderta* (South Eastern zone) and *Emba-alaje* district from Southern zone. Therefore, in this study, all the seven surveyed districts were purposively included in the first stage sampling. In the next stage, utilizing data from both Central statistics Agency (ECSA, 2012) [25], cluster *Tabias* or Enumeration Areas (EAs) were selected using probability proportionate to size (PPS). Accordingly, Enumeration Areas (EAs) from *Degua Temben* (n=4), *Emba-alaje* (n=4), *Enderta* (n=2), *Ganta Afeshum* (n=2), *Gulomekeda* (n=2), *Hawzen* (n=3), and *Saesie Tsaeda-emba* (n=4) were included. Larger population size *Tabias* or Enumeration Areas (EAs, n=21) were included so as to reach the number of households (HHs, n=25) required to achieve the overall sample size [(21 clusters ∈25 households) = 524 HHs] needed in this study (Figure 1). Then, a simple random sampling was used to select households and finally, the most knowledgeable family member or the head of a household (one individual per household) was approached for interview. Both male and female adults aged 18 years and above were eligible in this study.

**Figure 1:**
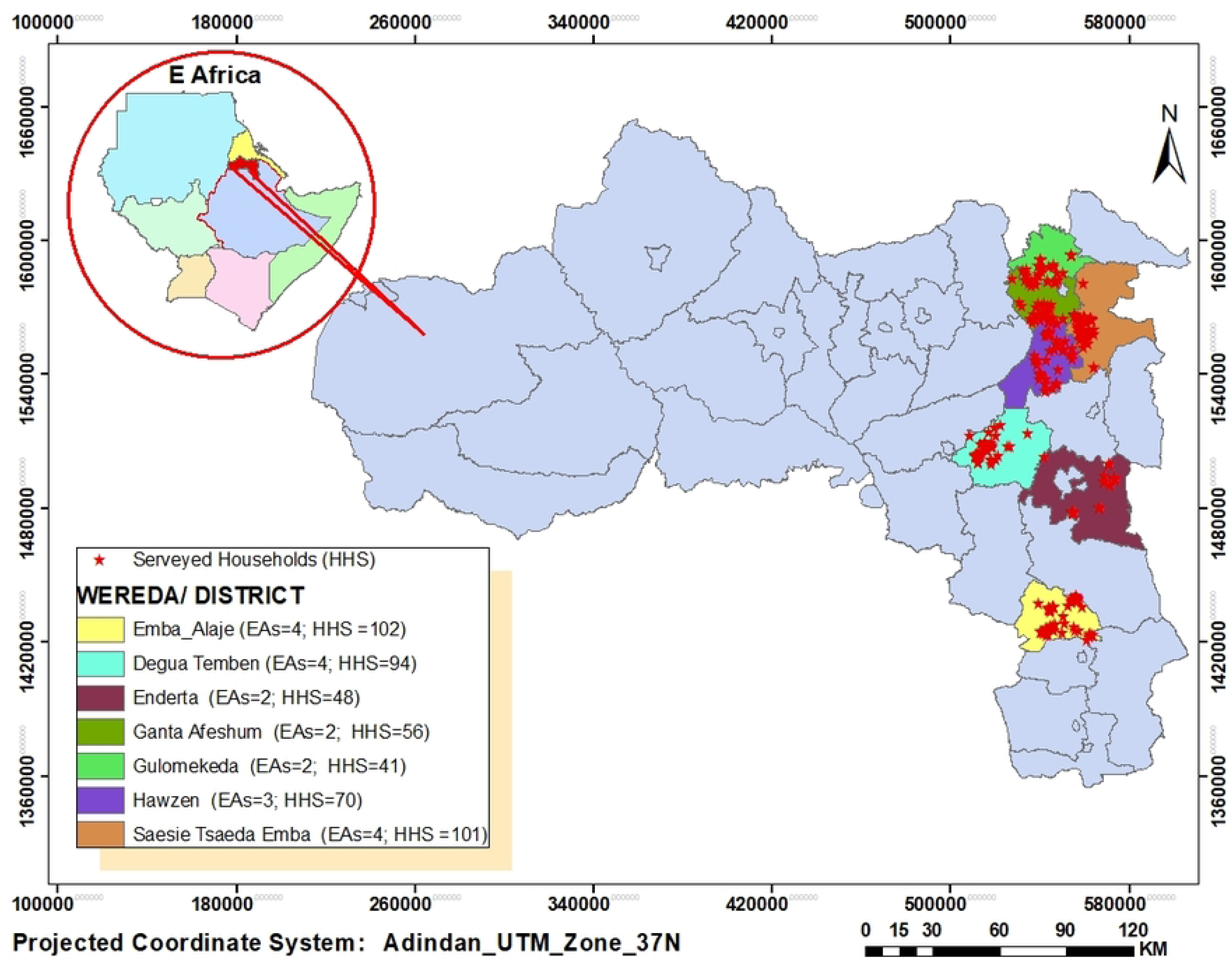
Study districts, number of Enumeration Areas (EAs) and number of surveyed households (HHs). The base map used is obtained from an openly available source at- https://open.africa/dataset/ethiopia-shapefiles

### Data collection tools and procedures

Data were collected using a structured questionnaire adopted from similar study literatures [15, 17, 21], pre-tested on 15 individuals recruited from households in the seven study districts. The questionnaire was designed to obtain information on participants’ socio-demographic characteristics; knowledge, attitude and practices (KAP); and treatment seeking behavior towards CL. The questionnaire was developed in English and then translated into the participants’ native language (Tigrigna), and the local name for CL (*Guzwa,* ጉዝዋ) was used to refer to the disease. The questionnaire was comprised of four sections.

<u>Section 1</u> of the questionnaire consisted of information on participants’ socio-demographic characteristics. <u>Section 2</u> contained questions on knowledge about CL that were designed to identify the primary sources of information used, to understand participants’ ability to identify the disease and explore their knowledge of aspects of CL, including signs and symptoms, local name of the disease, transmission mechanisms, treatment options, modern healthcare facilities, the existence of hyrax (the main reservoir host) and its relation with CL, and possible prevention measures for CL. <u>Section 3</u> contained questions designed to determine the participants’ attitudes towards CL as a health concern, stigma, whether the disease is curable by treatment, feelings on seeing people with CL, primary care provider choices and attitudes towards use of biomedical treatment for CL. <u>Section 4</u> consisted of questions designed to evaluate the treatment-seeking and prevention practices of the participants, such as primary healthcare options for active CL, reasons to choose a care provider and distance from healthcare facilities. Additional questions were included to assess potential risk factors, such as: occupation, time spent working outside (including overnight stays), use of hyrax dung as fertilizer, outdoor sleeping habits and preventive measures.

Questionnaires were administrated by a team of trained and experienced senior health professional data collectors who had previously been involved in leishmaniasis related household surveys. The data collection team performed house-to-house visits and were guided by community gatekeepers. When the head of household was not available, any responsible adult above 18 years selected by the family as the most knowledgeable person of the household was approached by the survey team for participation in the survey.

### Scoring methods

Scores for the KAP of respondents were developed using methods described in previous studies [9, 15, 20]. In brief, from the KAP questionnaire, a composite score of each question or item was calculated for each participant where each correct response was assigned a score of 1 and each incorrect or unsure response was assigned a score of 0. The total scores were further dichotomized based on the overall scores of each item. The total knowledge scores ranged from 0 to 9, and scores between 0 and 4 were categorized as poor knowledge, while scores between 5 and 9 were considered indicative of good knowledge. Similarly, the total attitude scores ranged from 0 to 6, scores between 0 and 3 were categorized as negative attitude, while scores between 4 and 6 were considered to denote a positive attitude. With regards to treatment-seeking behavior and prevention practices, scores were ranged from 0 to 12. Scores between 0 and 6 were considered to indicate poor prevention practices, while scores between 7 and 12 considered to be good prevention practices.

### Data analysis

The collected data were entered into EPI Info version 20.1.14 statistical package and exported to SPSS version 25.0 (SPSS, IBM Inc., Chicago) for statistical analysis. Descriptive statistics such as frequency, percentage and mean (standard deviation) were used, where applicable, to describe the KAP components with the explanatory (independent) variables. A chi-square test was used to examine the associations between KAP scores and the explanatory variables such as study area settings, age, gender, education level, occupation, and CL infection episode in a household. A multivariate logistic regression analysis was performed to identify the significant predictors of good knowledge on CL, positive attitude and good practices towards CL treatment-seeking behavior and prevention. Variables with a P value of ≤0.25 in the chi-square test were included in the logistic regression analyses. Adjusted odd ratios (AORs) and their corresponding 95% confidence intervals (CIs) were calculated based on the final models. The significance level for all tests was set at P < 0.05.

### Ethical considerations

Ethical clearance (Ref: ERC 1396/2019 and renewed on 15^th^ November, 2021; MU-IRB 2098/2021) was obtained from the Research Ethics Committee and Institutional Review Board (IRB) of the College of Health Sciences of Mekelle University and a permit letter (Ref: 882/1418/11) was granted by the Tigray Health Bureau (THB). Permissions to conduct the study in all of the study localities were obtained from each district health administrative offices. The purpose and the objectives of the study were clearly explained to all study participants to a level that they comprehended and verbal informed consent was obtained from all adult participants during the door-to-door interviews. During our door-to-door visits, all cases found suspected for non-severe active CL were advised to seek early diagnosis at CL care provider public hospitals in the region. While, those suspected for severe LCL, MCL and DCL cases were referred to Ayder comprehensive specialized hospital and linked with dermatology department staff of the hospital.

## Results

### Socio-demographic characteristics of the study participants

A total of 512 individuals (52.3% male and 47.7% female) participated in the study with 97.7% response rate. Of these, 70.5% were between 18 and 40 years old. More than half (52.3%) of the participants were from the eastern zone, a majority were rural inhabitants, and over three quarters (79.3%) were farmers by occupation. With respect to education level, 69.9% of the participants had attended modern schools and 30.1% of them had not received any formal education. Of the 358 educated participants, 39.4% and 29.1% had completed primary (1-6^th^ grades) and secondary (7-10^th^ grades) school levels, respectively. Only 7 (1.4%) respondents had completed college and university education level. From all participants, about 61% acknowledged that one or more CL episodes (at least one case identified through symptoms or diagnosed at health care) had occurred within their household during the period of the last five years (Table 1), showing that this disease is highly endemic in the region.

**Table 1:** Sociodemographic characteristics of study participants, Tigray, North Ethiopia (n=512)

| Variable | Categories | Frequency (n) | Percent (%) |
| --- | --- | --- | --- |
| Zone | Eastern | 268 | 52.3 |
|  | South eastern | 142 | 27.7 |
|  | Southern | 102 | 20.0 |
| Districts | Degua Temben | 94 | 18.4 |
|  | Emba-alaje | 102 | 19.9 |
|  | Enderta | 48 | 9.4 |
|  | Ganta Afeshum | 56 | 10.9 |
|  | Gulomekeda | 41 | 8.0 |
|  | Hawzen | 70 | 13.7 |
|  | Saesie Tsaeda Emba | 101 | 19.7 |
| Household setting | Rural | 375 | 73.2 |
|  | Urban/semi-urban | 137 | 26.8 |
| Age group | 18-40 | 361 | 70.5 |
|  | ≥41 | 151 | 29.5 |
| Gender | Male | 268 | 52.3 |
|  | Female | 244 | 47.7 |
| Educational background | Formal school unattended | 154 | 30.1 |
|  | Primary school (1- 6) | 202 | 39.4 |
|  | Secondary school (7-10) | 149 | 29.1 |
|  | College & above | 7 | 1.4 |
| Primary occupation | Daily laborer | 34 | 6.6 |
|  | Farmer | 406 | 79.3 |
|  | Trade/merchant | 26 | 5.1 |
|  | Government employee | 20 | 3.9 |
|  | Student | 26 | 5.1 |
| CL episode in household in the last five years? | Yes | 312 | 60.9 |
|  | No | 200 | 39.1 |

### Knowledge about cutaneous leishmaniasis

In the present study, most participants 97.3% (498/512) had some prior knowledge about the disease. The main sources of information around CL were household family members who had experience of the disease, and neighbours. Most participants (96.8%) described the disease using its local name “*Guzwa*” or ጉዝዋ) (the local name for CL, in Tigrigna language), while only 16 (3.2%) respondents had heard of the term ‘leishmaniasis’, the scientific name for the disease. A majority of the participants (71.5%) responded that skin lesion were the main sign of CL and over 95% described that the lesions appear on the face (Table 2).

**Table 2:**
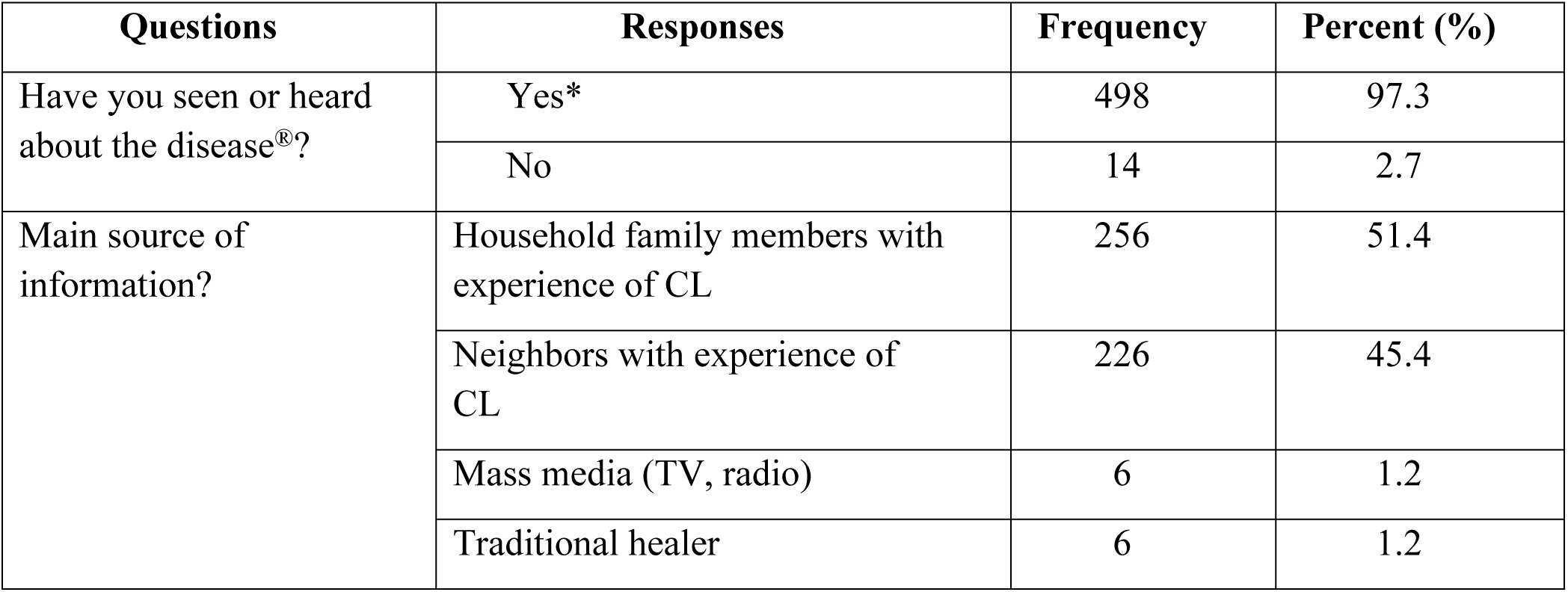
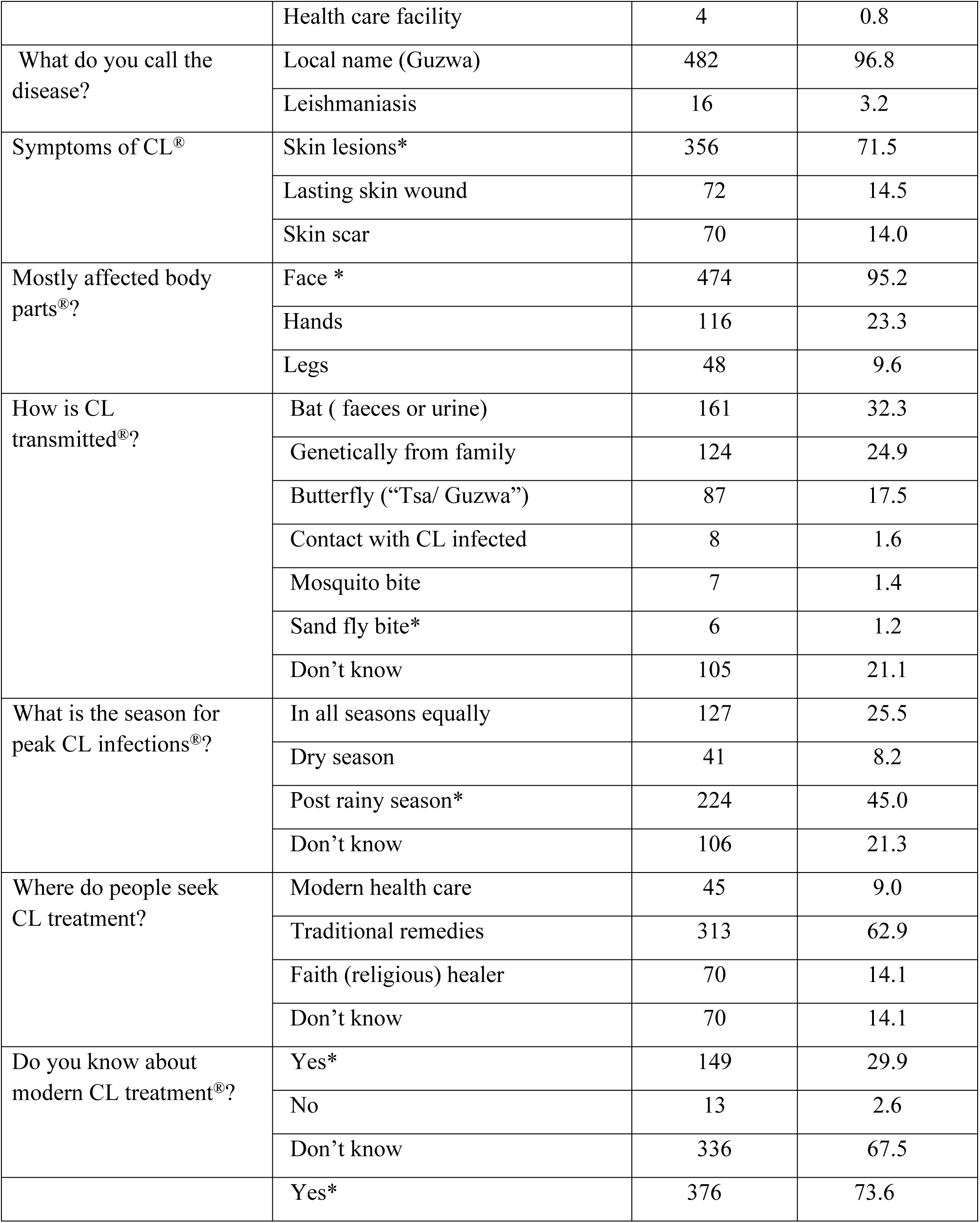
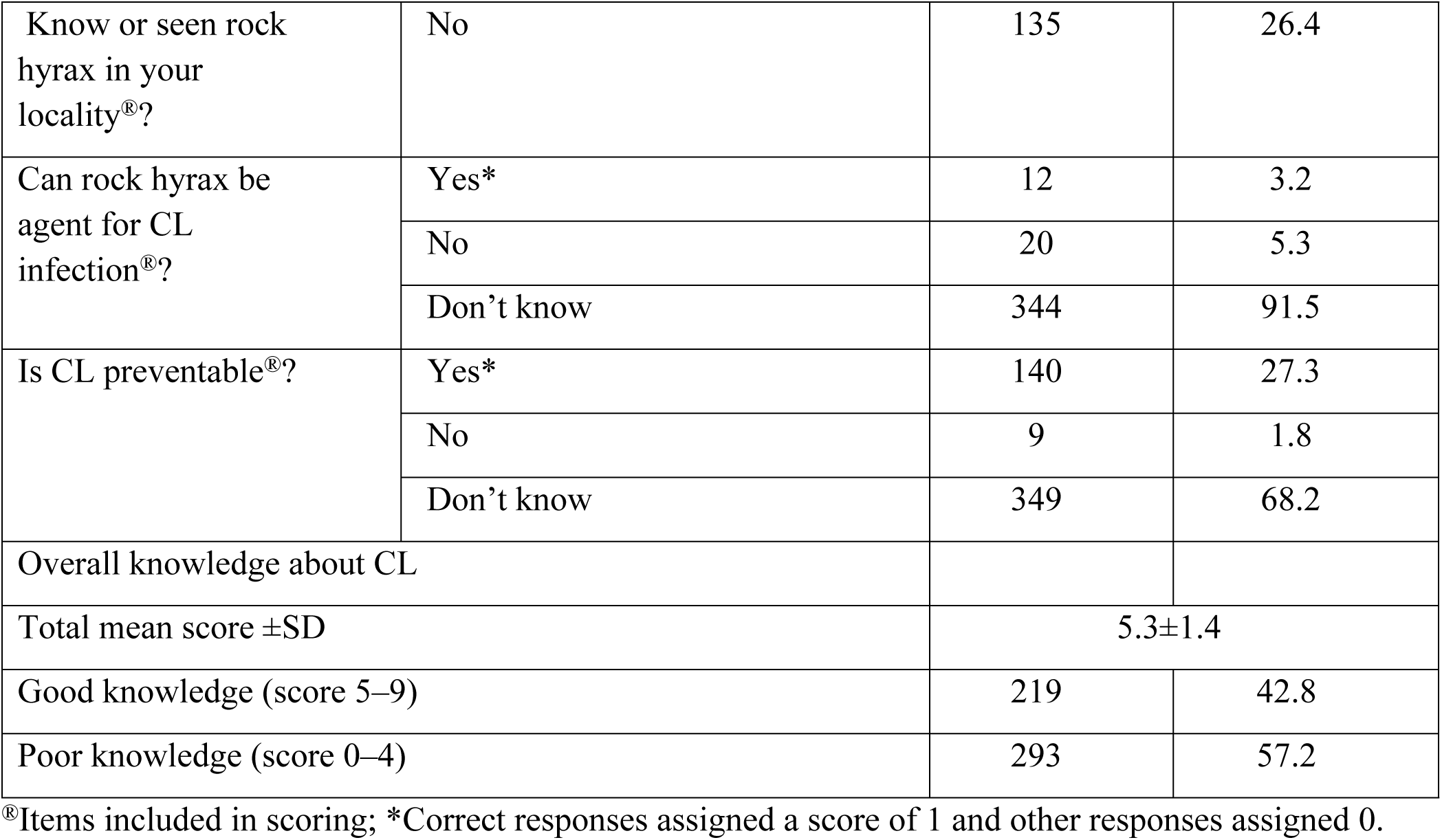
Knowledge about cutaneous leishmaniasis (CL), clinical presentations, its transmission, vector and reservoir hosts among communities in seven disease-endemic districts of Tigray region, 2022.

In this study, very few participants knew the biological cause and transmission route for CL. Only 6 (1.2%) participants were able to name the sand fly vector and 21.1% didn’t know any transmission mechanism. While 32.3% and 17.5% of the participants named bats, moths or butterflies as being disease vectors, respectively, 24.9% believed that CL was a genetically acquired disease. A majority of participants (73.6%) had seen rock hyrax in their localities, however, only 12 respondents (3.2%) knew that hyrax was a reservoir host for CL.

Regarding the peak season for CL infection, 224 (45.0 %) participants recognized that active wounds appear during the post-rainy season or immediately following summer which is the major rainy season in Ethiopia. In response to questions on treatment priorities, traditional healers (herbalist) was the primary choice for the majority (62.9%) of participants followed by religious healers/faith remedies (14.1%). Over 67% of the participants did not know about biomedical treatments for CL and about two thirds (68.2%) did not know about any CL prevention measures. Based on the overall scoring analysis, 219 (42.8%) participants had a good level of knowledge about CL (with scores of 5-9) while 293 (57.2%) had a poor level of knowledge (scores of 0-4) towards the disease (Table 2).

### Attitudes towards CL, its treatment and care provider priorities

Most participants (96%) perceived CL to be a serious public health concern in their locality. Over 67% of the participants responded that CL is a stigmatizing disease, while 150 (30.1%) participants didn’t know about CL-associated stigma and 13 (2.6%) participants responded that CL is not a stigmatizing disease. However, when participants were asked a subsequent question about their personal feelings when meeting people with a CL lesion, most of the participants (87.1%) responded that they felt uncomfortable.

A majority (87.8%) of participants responded that CL is a curable disease when treatment is given. Traditional healers (herbalists) were the primary choices for 73.6% of the participants and faith healers (religious remedies) were named as the first choice by 70 (16.4%). Modern healthcare (hospitals and clinics) was the first point of contact for only 45 (10%) respondents. Furthermore, only 29.9% of the participants responded that they were willing to seek modern healthcare facilities for CL treatment in the future. Based on the scoring analysis, 187 (36.5%) participants had a positive attitude toward CL health concern, stigma and discrimination, treatment and care provider priorities while 325 (63.5 %) had a negative attitude (Table 3).

**Table 3:**
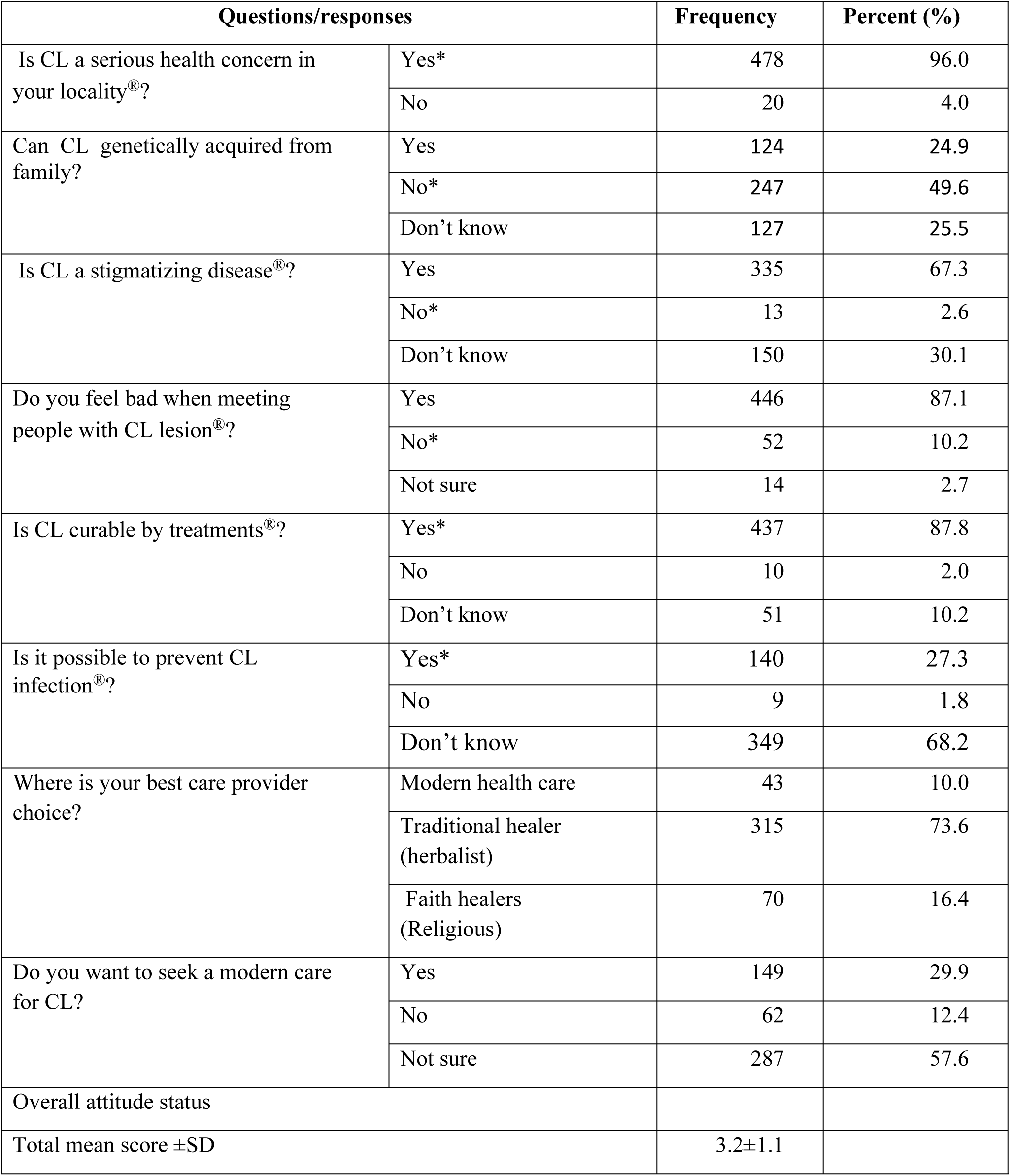
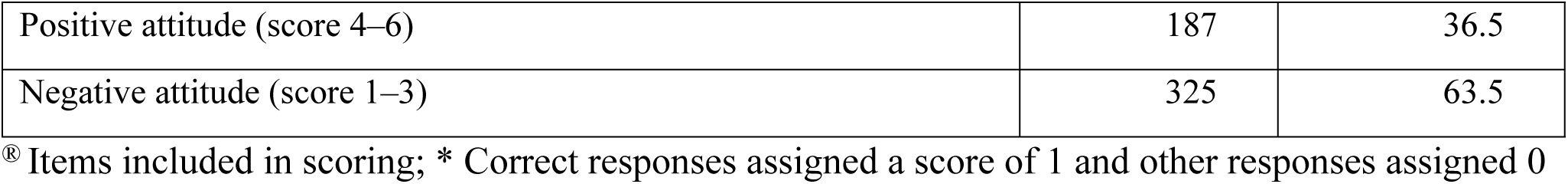
Attitudes toward cutaneous leishmaniasis (CL), its treatment and care provider priorities of community members in seven districts of Tigray region, northern Ethiopia, 2022.

| Questions/responses |  | Frequency | Percent (%) |
| --- | --- | --- | --- |
| Is CL a serious health concern in your locality<sup>®</sup>? | Yes* | 478 | 96.0 |
|  | No | 20 | 4.0 |
| Can CL genetically acquired from family? | Yes | 124 | 24.9 |
|  | No* | 247 | 49.6 |
|  | Don't know | 127 | 25.5 |
| Is CL a stigmatizing disease<sup>®</sup>? | Yes | 335 | 67.3 |
|  | No* | 13 | 2.6 |
|  | Don't know | 150 | 30.1 |
| Do you feel bad when meeting people with CL lesion<sup>®</sup>? | Yes | 446 | 87.1 |
|  | No* | 52 | 10.2 |
|  | Not sure | 14 | 2.7 |
| Is CL curable by treatments<sup>®</sup>? | Yes* | 437 | 87.8 |
|  | No | 10 | 2.0 |
|  | Don't know | 51 | 10.2 |
| Is it possible to prevent CL infection<sup>®</sup>? | Yes* | 140 | 27.3 |
|  | No | 9 | 1.8 |
|  | Don't know | 349 | 68.2 |
| Where is your best care provider choice? | Modern health care | 43 | 10.0 |
|  | Traditional healer (herbalist) | 315 | 73.6 |
|  | Faith healers (Religious) | 70 | 16.4 |
| Do you want to seek a modern care for CL? | Yes | 149 | 29.9 |
|  | No | 62 | 12.4 |
|  | Not sure | 287 | 57.6 |
| Overall attitude status |  |  |  |
| Total mean score ±SD | | 3.2±1.1 | |

|  |  |  |
| --- | --- | --- |
| Positive attitude (score 4–6) | 187 | 36.5 |
| Negative attitude (score 1–3) | 325 | 63.5 |

® Items included in scoring; \* Correct responses assigned a score of 1 and other responses assigned 0

### Treatment-seeking behaviors and practices

Table 4 shows responses around treatment-seeking behaviors and practices. When participants were asked about primary care options for active CL treatment in their community, most participants, (83%) stated that people used homemade herbal remedies as the primary method of care, while 63.3% used traditional remedies from a healer (herbalist), 13.7% used religious healing (faith remedies) and 17.4% of the participants used cauterization of lesions using very hot metal. The main reasons for selecting the primary care provider were proximity to home (48.8%) and perceived (good) reputation of the provider (29.7%).

**Table 4:**
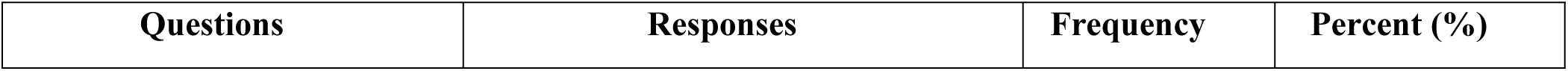
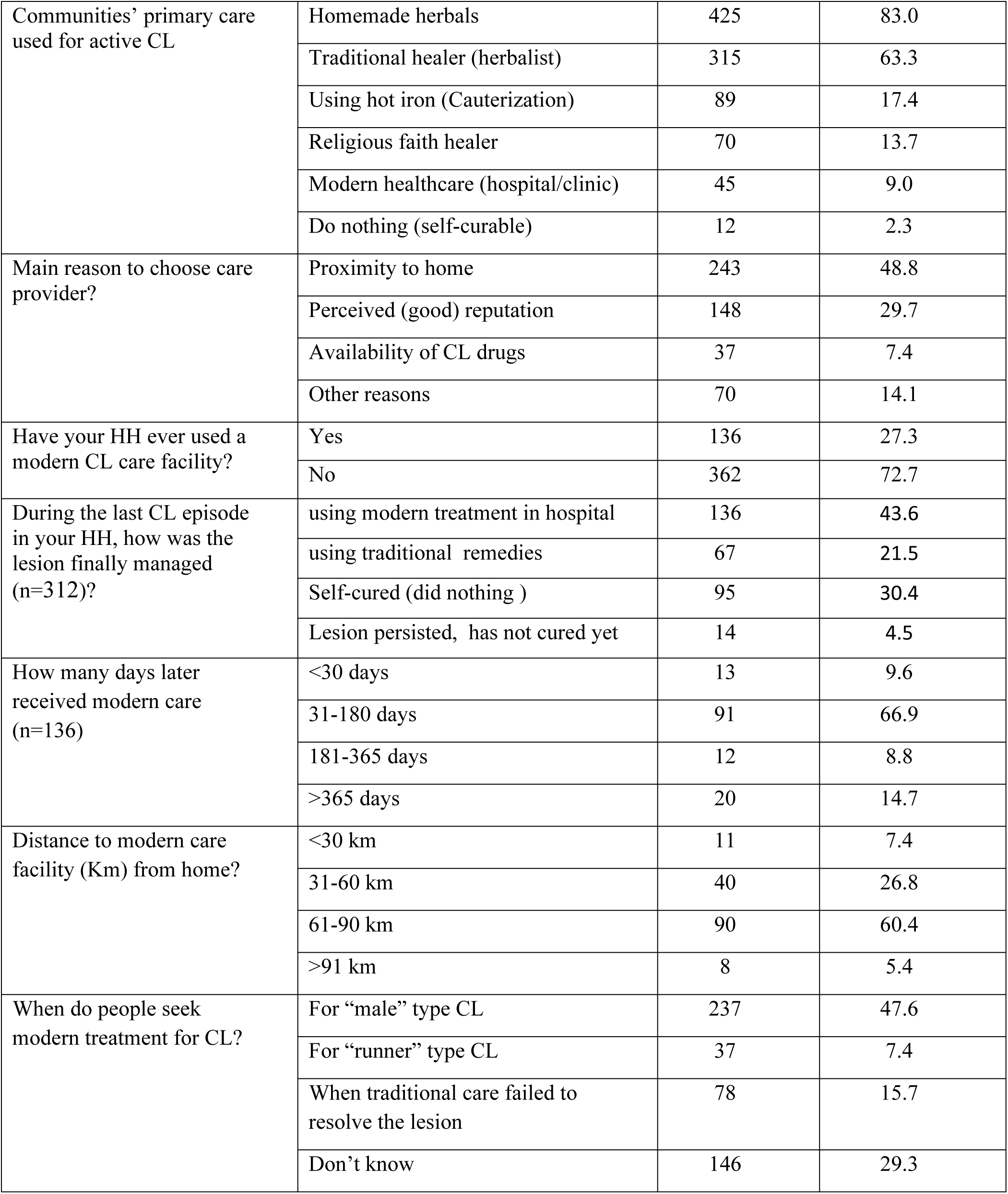
Treatment-seeking behaviors and practices of community members in seven districts of Tigray region, northern Ethiopia, 2022. (n=512)

Of all study participants, about 61% acknowledged that one or more CL episodes had occurred within their household during the last five years (Table 1). Of those, 43.6% of the participants approved that CL lesion had been treated and cured using modern care, 30.4% stated that lesion had cured by itself (without treatment), 21.5% used traditional remedies to cure lesion and 4.5% of the participants acknowledged that the lesion on household family has not cured yet (still persisting) (Table 4). About 27.3% (136/498) of the participants stated that they had used CL treatments at modern health care facilities, either for themselves or members of their household/families. Of those (n=136) who had used a modern CL care facility, about 85% had used one or more traditional remedies before receiving a treatment in modern health care facility, 66.9% had delays of 1-6 months and 14.7% had delays longer than 6 months before receiving this form of treatment. Regarding the nearest modern CL care facility, 60.4% of participants stated this was 61-90 km away from home and 26.8% responded 30-60 km away.

Participants were asked about situations where people prefer to seek modern healthcare for CL treatment; interestingly, this question revealed a concept of gendered lesions: 237 participants (47.6%) stated that they would seek healthcare when the CL lesion is “male” type, wording used to describe a more severe type of lesion that does not heal easily. Another 37 participants (7.4%) said they would seek modern healthcare when the CL is a “runner” (in Tigrigna, “Goyayit”) type, used to describe dissemination of the lesions, and 78 (15.7%) indicated they would seek healthcare when the wound failed to heal using traditional treatments (Table 4). Based on the scoring analysis outcome, while 173 participants (33.8%) had a good level of treatment seeking behavior and practices towards CL, 339 (66.2%) had a poor level of practices (Table 5).

**Table 5:** Common activities and prevention practices of community members in seven districts of Tigray region, northern Ethiopia, 2022 (n=512)

|  |  |  |  |
| --- | --- | --- | --- |
| Field work habit in both day and night? | Yes | 118 | 23.1 |
|  | No* | 394 | 76.9 |
| Morning up-to late evening field work habit? | Yes | 232 | 45.3 |
|  | No* | 280 | 54.7 |
| Outdoor / field/ sleeping habit | Yes | 298 | 58.2 |
|  | No* | 214 | 41.8 |
| Use of hyrax dung as fertilizer | Yes | 194 | 37.9 |
|  | No* | 318 | 62.1 |
| Did your HH ever received a modern CL care? | Yes* | 136 | 27.8 |
|  | No | 353 | 72.2 |
| Before receiving a modern CL care, did your HH used other care options (n=136)? | Yes | 115 | 84.6 |
|  | No* | 21 | 15.4 |
| Use prevention methods (n=489)? | Yes* | 140 | 28.6 |
|  | No | 349 | 71.4 |
| Sleep under a bed net | Yes* | 138 | 28.2 |
|  | No | 351 | 71.8 |
| Environmental hygiene | Yes* | 52 | 10.6 |
|  | No | 437 | 89.4 |
| Wear long sleeves/trousers | Yes* | 36 | 7.4 |
|  | No | 453 | 92.6 |
| Insecticide spray in house | Yes* | 34 | 7.0 |
|  | No | 455 | 93.0 |
| Use of insect repellents | Yes* | 11 | 2.2 |
|  | No | 478 | 97.8 |
| Overall treatment-seeking behaviors and prevention practices level |  |  |  |
| Total mean score ±SD | – | 4.48±2.02 | |
| Good (score 7–12) | – | 173 | 33.8 |
| Poor (score 0–6) | – | 339 | 66.2 |

\* Correct responses assigned a score of 1 and other responses assigned 0

Some practices were noticed in participants in the current study which may increase risk of *Leishmania* infection. Almost half, (45.3%) of the participants reported that they were commonly engaged working in fields from early morning until late evening, and 23.1% of the participants worked in fields during both daytime and at night, potentially increasing the risk of sand fly bites. Over half (58.2%) of the participants had outdoor sleeping habits and over one third (37.9%) of them used hyrax dung as fertilizer, potentially attracting sand flies.

Over 71% of the participants didn’t apply any known CL preventive measures, however, about 28.6% (140/489) of the participants stated one or more mechanisms. Of those, 28%, 10.6% and 7.4% of the participants mentioned using bed nets, improving environmental hygiene, and wearing long sleeves/trousers, respectively, as preventive measures for the disease. Based on the scoring analysis outcome, while 173 (33.8%) participants had a good level of prevention practices towards CL, 339 (66.2%) had a poor level of prevention practices (Table 5).

### Factors associated with knowledge about CL

Table 6 indicates both bivariate and multivariate logistic regression analyses results. Males were less likely to have good knowledge of CL as compared to female counterparts (p=0.004). Likewise, farmers were less likely to have good knowledge as compared to those who were non-farmers (e.g. students and employees) in occupation (p=0.021). Rural dwelling participants were 1.6 times more likely to have good level of knowledge towards CL compared to those living in urban and semi-urban resident areas(p=0.049). Participants who lived in a household with a previous CL case were over ten times (P<0.001) more likely to have a good knowledge of the disease relative to those dwelling in households with no previous CL episodes.

**Table 6:** Demographic factors compared with participants’ knowledge about cutaneous leishmaniasis (CL), in Tigray, northern Ethiopia, 2022.

| Variables and categories | Knowledge about CL |  | COR (95% CI) | AOR (95% CI) | P value<br>AOR |
| --- | --- | --- | --- | --- | --- |
|  | Good | Poor |  |  |  |
| Gender |  |  |  |  |  |
| Male | 124 (46.3%) | 144(53.7%) | 0.74 (0.52, 1.05) | 0.55(0.36, 0.83) | 0.004 |
| Female | 95(38.9%) | 149(61.1%) | 1 | 1 |  |
| Age group (year) |  |  |  |  |  |
| 18-40 | 155(42.9%) | 206(57.1%) | 1.02 (0.70, 1.50) | 0.85(0.52, 1.38) | 0.507 |
| >40 | 64(42.4%) | 87(57.6%) | 1 | 1 |  |
| Educational background |  |  |  |  |  |
| Educated | 70(45.5%) | 84(54.5%) | 1.17 (0.80, 1.71) | 0.79 (0.48, 1.29) | 0.354 |
| Formal school<br>unattended | 149(41.6%) | 209(58.4%) | 1 | 1 |  |
| Primary occupation |  |  |  |  |  |
| Farmer | 183(45.1%) | 223(54.9%) | 1.60 (1.02, 2.50) | 0.53 (0.31, 0.91) | 0.021 |
| Non-farmer | 36(34.0%) | 70(66.0%) | 1 | 1 |  |
| Household setting |  |  |  |  |  |
| Rural | 155(41.3%) | 220(58.7%) | 0.80 (0.54, 1.19) | 1.60 (1.00, 2.57) | 0.049 |
| Urban or semi-urban | 64(46.7%) | 73(53.3%) | 1 | 1 |  |
| Previous or current CL episode in household |  |  |  |  |  |
| Yes | 190(69.9%) | 122(39.1%) | 0.11 (0.07, 0.17) | 10.19(6.36,16.30) | <0.001 |
| No | 29(14.5%) | 171(85.5%) | 1 | 1 |  |

### Factors associated with participants’ attitude towards CL

These results showed that participants’ occupation and household settings were significant factors associated with attitude towards CL disease (P<0.05). Participants who engaged in farming activities as their primary occupation were 60% times more likely to have a negative attitude towards CL compared to those who were not farmers (e.g. students and employees) in occupation. Participants from urban and semi-urban areas were about 2.2 times (P<0.001) more likely to have positive attitude related to CL infections, exclusion and stigmatisation of infected individuals compared to those rural-dwelling counterparts (Table 7).

**Table 7:** Factors associated with study participants’ attitudes towards cutaneous leishmaniasis (CL), among community members in seven districts of Tigray region, northern Ethiopia, 2022.

| Variables and categories | Attitude about CL |  | COR (95% CI) | AOR (95% CI) | P value for AOR |
| --- | --- | --- | --- | --- | --- |
|  | Negative | Positive |  |  |  |
| Gender |  |  |  |  |  |
| Male | 92 (34.3%) | 176(65.7%) | 1.22 (0.85, 1.77) | 1.18 (0.81, 1.71) | 0.392 |
| Female | 95(38.9%) | 149(61.1%) | 1 | 1 |  |
| Age group (year) |  |  |  |  |  |
| 18-40 | 136 (37.7%) | 225 (62.3%) | 1.18 (0.79, 1.76) | 0.73 (0.47, 1.14) | 0.165 |
| >40 | 51(33.8%) | 100(66.2%) | 1 | 1 |  |
| Educational background |  |  |  |  |  |
| Educated | 128 (35.8%) | 230 (64.2%) | 1.12 (0.75, 1.65) | 0.79(0.50, 1.24) | 0.310 |
| Formal school unattended | 59(38.3%) | 95 (61.7%) | 1 | 1 |  |
| Primary occupation |  |  |  |  |  |
| Farmer | 155(38.2%) | 251(61.8%) | 1.43 (0.90, 2.26) | 0.60 (0.36, 0.99) | 0.048 |
| Non-farmer | 32(30.2%) | 74(69.8%) | 1 | 1 |  |
| Household setting |  |  |  |  |  |
| Rural | 122(32.5%) | 253(67.5%) | 0.53 (0.36, 0.80) | 2.17 (1.42, 3.31) | <0.001 |
| Urban/semi-urban | 65(47.4%) | 72(52.6%) | 1 | 1 |  |
| Previous or current CL episode in household |  |  |  |  |  |
| Yes | 123(39.4%) | 189(60.6%) | 0.72 (0.50, 1.05) | 1.37 (0.94,2.01) | 0.102 |
| No | 64(32.0%) | 136(68.0%) | 1 | 1 |  |

### Factors associated with treatment-seeking and prevention practices

Our findings showed that gender and previous/current CL episodes in the household were significantly associated factors with treatment seeking behavior and prevention practices (P<0.05). While male participants were about 1.5 times (P=0.044) more likely to follow poor treatment-seeking and prevention practices compared to female counterparts, educated participants were 39% times less likely to have poor practices compared to those participants with no formal education (P<0.001). The frequency of good treatment-seeking and prevention practices was also increased by about 3 times (P<0.001) among participants who lived in households with previous/current CL cases compared to those households with no previous CL episodes (Table 8).

**Table 8:** Factors associated with participants’ treatment-seeking and prevention practices for cutaneous leishmaniasis (CL), in Tigray region, northern Ethiopia, 2022.

| Variables and categories | Treatment-seeking and prevention practices |  | COR (95% CI) | AOR (95% CI) | P value for AOR |
| --- | --- | --- | --- | --- | --- |
|  | Good | Poor |  |  |  |
| Gender |  |  |  |  |  |
| Male | 79(29.5%) | 189(70.5%) | 1.22 (0.85, 1.75) | 1.49 (1.01, 2.22) | 0.044 |
| Female | 94(38.5%) | 150(61.5%) | 1 | 1 |  |
| Age group (year) |  |  |  |  |  |
| 18-40 | 112(31.0%) | 249 (69.0%) | 1.18 (0.79, 1.76) | 1.01 (0.63, 1.59) | 0.982 |
| >40 | 61(40.4%) | 90(59.6%) | 1 | 1 |  |
| Educational background |  |  |  |  |  |
| Educated | 73(47.4%) | 81(52.6%) | 1.12 (0.75, 1.65) | 0.39 (0.24, 0.62) | <0.001 |
| Formal school unattended | 100(27.9%) | 258(72.1%) | 1 | 1 |  |
| Primary occupation |  |  |  |  |  |
| Farmer | 139(34.2%) | 267(65.8%) | 1.43 (0.90, 2.26) | 1.52 (0.91, 2.54) | 0.112 |
| Non-farmer | 34(32.1%) | 72(67.9%) | 1 | 1 |  |
| Household setting |  |  |  |  |  |
| Rural | 136(36.3%) | 239(63.7%) | 0.53 (0.36, 0.80) | 0.67 (0.42, 1.06) | 0.086 |
| Urban/semi-urban | 37(27.0%) | 100(73.0%) | 1 | 1 |  |
| Previous/current CL episode in household |  |  |  |  |  |
| Yes | 132(42.3%) | 180(57.7%) | 0.72 (0.50, 1.05) | 2.99 (1.96, 4.57) | <0.001 |
| No | 41(20.5%) | 159(79.5%) | 1 | 1 |  |

## Discussion

This study evaluated the knowledge, attitude and practices (KAP), treatment seeking behaviour and disease prevention practices related to cutaneous leishmaniasis (CL) among communities located in seven disease-endemic districts of Tigray region, northern Ethiopia. CL was notable among the current study communities and perceived to be a serious public health challenge. About 61% of the participants also stated that one or more CL episodes had occurred within their household during the last five years, showing that this disease is highly endemic in the region. The findings revealed that the study participants had poor level of knowledge related to CL transmission and poor level of attitudes towards people with CL lesion, where over 67% of the participants believed that CL is a stigmatizing disease and over 87% acknowledged that they felt uncomfortable when meeting people with active lesion, indicating that CL stigma is common. Besides, the study findings shown that participants had poor level of treatment seeking and prevention practices. The participants’ overall poor level of KAP and treatment seeking behavior and prevention practices revealed in this study could most likely be deeply rooted in the neglected nature of the disease in government health policies and a lack of information on disease control measures.

In this study, overall, 42.8% of the participants had good level of knowledge about cutaneous leishmaniasis. This finding was higher than studies carried out in Delanta district, Northeast Ethiopia [26], in Wolaita zone, Southern Ethiopia [20], Kutaber District, Northeast Ethiopia [22] and in Shara’b district, Southwestern Yemen [15]. However, the current result was lower than studies conducted in Gamo Gofa zone, Southern Ethiopia [17] and a study among communities in hyperendemic areas in western Yemen [9]. This knowledge level discrepancy could be attributed to variations in magnitudes of CL prevalence among investigated countries, differences in specific studied area settings, and due to socio-demographic and sociocultural variations of each studied populations. However, the poor levels of knowledge towards CL revealed in this study could show adequate awareness creation activities about this disease were not in place and the neglected nature of the disease still continues in Ethiopia [5] including in the current study communities.

When we examine specifically beyond the overall knowledge level, most participants were familiar with the disease and described it using a common local term ‘Guzwa’ (a name for CL in Tigrigna language); while only very few had heard the scientific name of the disease, ‘leishmaniasis’. In the current study, 96.8% of participants had seen CL cases within the study communities, either from infected household family or neighbors, and considered a skin lesion on the face to be a symptom of CL. The participants’ high awareness of the signs and symptoms of CL is most likely due to the high prevalence of the disease in the study areas. Awareness of these aspects was higher than earlier studies carried out in Amhara region, Ethiopia, where 85.6% of participants had seen cases of CL in their localities [22]. Similarly, in the Volta region of Ghana and southwestern Yemen, 82.0% and 76% of the study participants, respectively had seen CL cases either among family members or other people in the study localities [15, 27]. However, in contrast with the present findings, in a KAP study carried out in a non-endemic area of Alexandria, Egypt, most participants (90%) had never seen CL infected person [28]. This difference is likely a reflection of variation in CL prevalence between these geographical regions.

In the current study, although most of the participants showed encouraging knowledge about symptoms of CL, they had an overall poor level of knowledge towards its transmission mechanisms, only six participants (1.2%) mentioned the sand fly vector by name. This finding is comparable with a similar KAP study carried out in Wolaita zone, southern Ethiopia, where none of the participants had heard of the sand fly [20]. The current finding is also consistent with other similar studies carried out in different regions of Ethiopia [17, 21]. Moreover, the current study participants had high level of misconceptions towards transmission mechanisms of the disease and the causal agents. About one quarter of the current participants considered CL to be a genetically acquired disease, about one third considered bats as being the disease vector and about one fifth of the participants believed that butterflies or moths could be responsible for spreading the disease. These findings are similar to previous studies conducted in Ethiopia. For example, 61.6% of study participants in Kutaber District had misconceptions on the mode of transmission [22], 19.5% of participants in a study in Delanta District responded that CL is transmitted via bat urine [26]. However, unlike to current study findings, a similar KAP study in central Iran found that 97.9% of the participants had a theoretical knowledge of the sand fly vector [29]. The differences could be due to variations in sociodemographic factors, including the study settings and education level of participants. The study in Iran was institution-based and conducted with student participants [29] whereas the current study was conducted in CL-endemic community settings where 30% of participants had not received formal education.

In overall, 36.5% of the current participants had a positive attitude towards CL infected individuals. This finding was comparable with earlier studies undertaken in Northeast Ethiopia and studies in western Yemen, where 34.5% and 38.1% of the participants respectively, had positive attitude towards people with CL infections [9, 26]. On the other hand, the present finding is higher than a study result found in Kutaber district of Ethiopia, where only 18.2% of the study participants had a positive attitude towards CL infections [22]. The discrepancy with later report could be due to variation in beliefs and cultural differences towards the disease, where there could be higher fear of CL infection among Kutaber district community relative to the study communities in Tigray. Besides, more than half of the participants in this study demonstrated a negative attitude towards people with CL infections. This could be due to lack of scientific information about the disease and poor access to CL treatment facilities. Several studies described that CL lesions and disfigurements have multifaceted psychosocial impacts on affected individuals [31] and the direct consequence of a negative attitude towards CL has been indicated to be a barrier for early treatment seeking behavior [12, 32]. While CL is often described as self-healing without drug treatment, this may take many months. Lack of treatment may lead to the development of more severe pathology and there is an increased risk of transmission to others in the community. Delays in treatment may lead to several irreversible complications due to secondary infections, which could result in deep tissue damage and disfiguring CL scars [33]. Several studies in Afghanistan, Brazil, Morocco and Yemen have indicated that, depending on the severity and visibility of CL disfigurements, most cases were experiencing some degree of psychosocial morbidities including reduced self-value or self-stigma, enacted stigma, social stigma and discrimination [34, 35].

Looking at perceptions of CL in the community, a large majority of participants (96%) responded that this disease is a serious public health concern in their locality. This finding is even higher than earlier studies undertaken in southern Ethiopia, northwest Ethiopia and western Yemen; where 71.6%, 82% and 85.1% of the participants respectively, responded that CL was a serious public health challenge [9, 20, 21] and likely reflects the high prevalence in the study areas and the effects on affected individuals and communities. In the current study, over 67% of the participants perceived CL to be a stigmatizing disease, which is higher than previous studies in southern Ethiopia and in western Yemen [9, 20]. The high level of stigma revealed in this study could be associated with poor understanding on the mode of disease transmission. For example, a considerable number of our participants believed that CL is a genetically acquired disease. The actual level of CL stigma among the study communities could be even higher than the current findings as when our participants were being asked a subsequent question about their feelings, 87.1% of participants acknowledged that they felt uncomfortable when looking at people with CL. The stigma associated with CL can result in social discrimination, isolation and rejection, and may also exacerbate health outcomes and influence the educational attainment of children [36–38]. Timely treatment and effective disease control measures are needed to minimise these wider effects of CL.

In this study, about 34% of participants had a good level of treatment-seeking behaviour and prevention practices. This finding is about equivalent to a similar study carried out in southern Ethiopia, where about 37.5% of the study participants had good levels of practices [17]. On the other hand, the present finding is higher than a recent KAP study undertaken in western Yemen, where only 16.3% of the study participants had good levels of treatment-seeking and prevention practices [9]. This discrepancy could be due to the prolonged civil war in Yemen, which brought the public health system to collapse [9, 39], which could be contributed for reduction of patients’ healthcare utilization status in the country.

In the current study, over 66% of the participants had a poor level of treatment-seeking behaviour. Similar to the war-crisis in Yemen, the war in the Tigray region of northern Ethiopia, erupted in November 2020, has brought unimaginable humanitarian crisis and enormous damage to the health system in the region [40]. As a result, the health system in Tigray has been almost collapsed; about 80% of the primary hospitals and 86% of the secondary and tertiary hospitals including the CL care provider hospitals were fully or partially damaged and/or vandalized/looted and hence the majority of the health facilities went non-functional. Those few in operation have been functioning at a much lower capacity due to the displacement, relocation, migration, injury and/or death of the health workforce and shortage of drugs, medical supplies and equipment [41]. Therefore, the war associated health system damage in Tigray could be one of the reasons for the poor level of treatment-seeking behaviour revealed among the vast majority of the current study participants.

In the present study, females had higher scores in questions relating to good CL treatment-seeking and prevention practices compared to males. This finding could be directly related to the greater psychosocial impact of CL among females relative to males. Several previous studies have indicated a higher psychosocial burden of CL found in women than in men [35]. Some studies also implicated that women with visible CL scars, mainly on the facial areas, were found to bear significant psychosocial burden including on their marriage opportunities [42], and were more likely to express a sense of shame and embarrassment of being seen in public [9]. In remote rural areas such as our study sites in Tigray, formal healthcare facilities offering CL diagnosis and treatment are located a long distance from CL-endemic regions, are difficult to access and require both time and monetary costs. The direct and indirect economic costs of healthcare for visceral leishmaniasis have previously been assessed in Tigray [41]. In male-dominated rural communities like Ethiopia [43], beyond embarrassment and self-stigmatization, economic constraints could provide additional challenges for females to access CL healthcare. Under these circumstances, the most feasible options may be to remain at home until skin lesions self-cure or to use traditional remedies, with the risk of more severe pathology.

Our findings showed that over 63% of participants preferred to seek remedies from traditional healers (herbalists) rather than formal CL healthcare facilities. This could be due to several factors, including local beliefs, lack of awareness about biomedical treatments available and poor access to facilities. These findings are consistent with many community-based studies in Ethiopia: traditional remedies were the primary choice for 67.6% of participants in Gamo-Gofa zone [17], 68.3% of participants in Amhara region [21] and about 77% of participants in Wolaita zone [20]. A study in Nigeria also found that participants preferred to seek CL treatments with traditional healers rather than formal healthcare services [44].

The current findings revealed that religious/faith healers (locally named *“Tsebel”*) offered traditional treatments specifically called *“Tsebel Senbet Senabti”*, which were among the most common traditional methods used to treat CL in the study sites. These findings are consistent with previous studies in southern Ethiopia [20]. Moreover, over 60% of the current participants acknowledged that the nearest formal CL healthcare facility is located approximately 61 to 90 kilometers away from home. Most formal CL healthcare in Ethiopia, including in Tigray, is centralised and located in regional or zonal capital cities [45], a long distance from endemic areas. In order to improve access, decentralization of CL treatment facilities to the disease endemic localities could be one of the crucial measures needed.

Our findings showed that most participants did not know about CL prevention mechanisms. A very small proportion stated that bed nets or mentioned different preventive methods. These findings are consistent with similar studies conducted in southern Ethiopia and western Yemen [9, 20], where most of the study participants had poor level of knowledge about the insect vector and its preventive measures. About three quarters of the current participants had seen rock hyraxes in their localities and over one third of them also responded that they used hyraxes’ dung as a fertilizer in their farm plots. Hyrax species have been shown to be the primary reservoir hosts of CL parasites in Ethiopia [46] and a study carried out in eastern Tigray has also indicated that the presence of hyraxes close to resident houses was significantly associated with CL infections [47]. However, only a very small number of our participants were informed regarding hyraxes as potentially involved in CL transmission. Considering the present findings, the communities living in CL endemic areas of Tigray do not have adequate information towards prevention and control of the disease. Providing more education on CL transmission, the insect vector (sandfly), the reservoir hosts (hyrax) and activities/times that are more risky would help communities to take preventive measures against the disease.

### Strengths and limitations of the study

This study included wider geographic coverage (seven districts from three zones) of Tigray; hence, the overall findings could represent the entire communities’ knowledge, attitude, treatment-seeking behavior and prevention practices towards CL in the region. Given the above strength, this study has the following limitations. First, this study was done in a single region, Tigray, that the findings may not be generalized to the overall population in Ethiopia. The second limitation could be related to the nature of cross-sectional study design used, which may influence participants for recall bias. The third limitation could be related to the participants’ responses for some questions, especially related to treatment-seeking behavior of the respondents, which may predispose them for social desirability biases. Hence, the findings of this study should be interpreted with caution.

### Conclusion and recommendations

In this study, more than one half of the study participants had poor level of knowledge about CL and nearly all participants didn’t know about the disease transmission mechanism, the insect vector (sand fly) and reservoir hosts. Besides, more than two-third of participants had unfavorable attitude towards the disease and majority (67%) perceived CL to be a stigmatizing disease. Moreover, about two-third of the participants had poor understanding towards CL treatment in formal healthcare facilities; instead, traditional methods were the primary choices for most participants. The majority of the participants had very low scores for questions around practices related to prevention and control of the disease. More than half of respondents didn’t apply any known prevention measures and a significant proportion of respondents didn’t know about any mechanism of CL prevention. Occupation and rural household settings were determinants significantly associated with participants’ attitude. While gender, education level and previous CL episode in the households were significant predictors associated with treatment seeking behavior and prevention practices. These findings are reflective of the absence of adequate awareness creations towards prevention and control efforts in disease-endemic areas of Tigray. This is in contrast to the perception of communities who responded that CL is a major health problem and is stigmatising to those individuals affected. These findings give strong support for an integrated intervention through community mobilization and health education campaigns in these disease-affected populations focusing on the CL causal agents, disease transmission mechanisms and its prevention measures.

## Data Availability

The datasets are available at the Chief Executive Director Office of College of Health Sciences of Mekelle University. Therefore, the data sets used in this article are available at: and would be provided on reasonable requests.

## Acknowledgements

The authors would like to acknowledge all participants who voluntarily accepted the data collection team and served as respondents in the study for their valuable time and information; the data collectors who contributed their invaluable time and energy during data collection and all those who in one way or another contributed their thoughts in this study are highly appreciated.

## Authors’ contributions

This report is part of a dissertation by SBT who conceived the project idea, wrote the draft proposal, applied for ethical clearance, collected data, analyzed data and wrote the manuscript. AMB and HPP supervised SBT during inception of the proposal, reviewed and edited the proposal, assisted in interpretation of the results and writing the manuscript. All three of the authors have critically read and reviewed and approved the manuscript for publication reviewed and edited the manuscript

## Availability of data and materials

The datasets used in this study are available from the corresponding author on reasonable request.

